# Perception of Safety in Behavioral Health Crisis Units among Patients and Care Partners versus Artificial Intelligence (AI): A Multimethod Study

**DOI:** 10.64898/2026.04.06.26350257

**Authors:** Roxana Jafarifiroozabadi

## Abstract

**Background:** Safety is a critical concern in behavioral health crisis units (BHCUs), where environmental risks (e.g., ligature points) can lead to injury to self or others. However, limited research has examined how perceived safety influences facility selection among patients and care partners, or how these perceptions align with AI-driven safety risk assessments in such environments.

**Method:** To address these gaps, a nationwide discrete choice online survey was conducted using image-based scenarios of BHCU environments, where participants selected preferred facilities based on a range of attributes, including environmental safety risks (e.g., ligature points). Additionally, participants identified safety risks in survey images, which were compared with outputs from an AI-driven tool developed and trained to detect environmental risks by experts. Quantitative analysis using conditional logit models examined the influence of attributes on facility choice, while spatial comparisons of annotated images and heatmaps assessed participant and AI-identified risk alignments.

**Results:** Findings revealed that the higher frequency of safety risks in images significantly reduced the likelihood of facility selection (p < .001, OR ≈ 1.28), highlighting the importance of perceived safety in user decision-making. While there was notable alignment between heatmaps generated by participants and AI, key differences emerged, suggesting that participants’ safety perception was influenced by features not fully captured by AI, such as the type of materials or unknown, out-of-label safety risks in facility images.

**Conclusions:** Despite these limitations, results highlighted the value of integrating AI-driven assistive tools for non-expert user safety risk assessment to support decision-making for safer BHCU environments.

## 1. Introduction

Safety remains a major concern in the design of mental and behavioral health environments, where challenges related to sentinel events, such as injury to self or others can contribute to significant distress among patients and staff (Norouzi, 2024; Oostermeijer et al., 2021; Ulrich et al., 2018). The built environment’s impact on safety experience and occurrence of such events in mental and behavioral health units is well documents in the existing literature. Environmental features such as enhanced privacy, decreased crowding, improved visibility, access to sources of positive distraction (e.g., views of nature or daylight), and family-like designs have been shown to shape patients’ experience and impact their well-being and outcomes in such units (Hunt et al., 2012; Papoulias et al., 2014; Thibaut et al., 2019; Weber et al., 2022). Traditionally, safety in the environment of care is defined, evaluated, and applied by experts (e.g., healthcare designers, medical planners, etc.) through standardized guidelines for health facilities design, such as Facility Guidelines Institute (Facility Guidelines Institute., 2022) or based on the input obtained from end-users and stakeholders during the design process of the facilities (Pankey et al., 2022; Sachs et al., 2020; Ulrich et al., 2018; US Department of Veterans Affairs, n.d.). In mental and behavioral health facilities, safety is also associated with features, such as ligature risks, defined as points where a cord, rope, bedsheet, or other fabric/material can be looped or tied to create a substantial point of attachment that may result in self-harm or loss of life (Hunt et al., 2012; Pankey et al., 2022; US Department of Veterans Affairs, n.d.).

Research suggests that safety in healthcare environments is not only a technical construct but also a perceptual one which may influence user’ willingness to engage with care (Listiowati et al., 2023). Nevertheless, there is a gap in the literature regarding how the perceived safety of the care environment can influence facility selection among users during mental and behavioral health crises. When selecting facilities for receiving care, users often rely on a range of factors, such as insurance coverage, accessibility and travel time, appointment availability, facility ratings, and quality of care based on facility ratings (Broek-Altenburg & Atherly, 2020; Schwartz et al., 2021; Smith et al., 2018; Woo et al., 2022). Prior research in healthcare decision-making suggests that environmental features and visual impressions can also shape patients’ choices of care settings (Woo et al., 2022). However, it is unclear to what extent mental and behavioral healthcare users, including patients and their care partners, understand environmental safety features (e.g., ligature risks) based on visual cues within the built environment and whether their perception of safety impacts their selection of mental and behavioral health facilities to receive care at the time of crisis.

On the other hand, recent advancements in artificial intelligence (AI) and computer vision systems have contributed to automatic detection of environmental safety risks (Ayoubi & Arashpour, 2026). For instance, AI-driven tools have been trained by experts and employed in healthcare settings to automatically detect adherence to protocols (e.g., personal protective equipment (PPE) or hand hygiene) and have shown equal or superior performance and sensitivity compared to human observers (Kim et al., 2025; Singh et al., 2020). However, there is still a notable gap in leveraging AI-driven assistive tools for safety assessment of the built environment features in mental and behavioral healthcare environments, such as the behavioral health crisis unit (BHCU), especially for non-experts, including stakeholders, patients, or care partners. To date, most stakeholders rely on their lived experience, personal knowledge, intuition, and contextual understanding when evaluating safety in environments at the time of crisis while AI-driven tools commonly perform based on expert data input and existing guidelines (Lebovitz et al., 2021).

This research addresses the gaps mentioned above and aims to understand, define, and compare the perception of safety between objective tools driven by AI and end-users (patients and their care partners) through addressing the following questions: 1) How does perception of safety (e.g., reduced ligature risks) impact facility selection among end-users when seeking care in a mental and behavioral health facility, such as the BHCU?, and 2) What would be the divergence and congruence in perception of safety risks in mental and behavioral health environments, such as the BHCU, between end-users and AI-driven tools? The study is particularly focused on understanding this relationship in the context of recently developed model of crisis care in BHCUs due to the need for immediate stabilization, intervention, and treatment within the first 24 hours of the patient’s first encounter with the facility before mental and behavioral symptoms exacerbate (Balfour & Carson, 2024; Balfour & Zeller, 2023; Jafarifiroozabadi et al., 2026). These environments commonly accommodate a spectrum of high-to-low risk patients with distress, loss of control, suicidal thoughts, self-harm or harming others, and therefore can significantly influence how safety is perceived, experienced, and understood.

## 2. Methodology

This study incorporated a multimethod approach to examine similarities and differences in the detection and perception of environmental safety risks between AI and users of mental and behavioral healthcare environments, such as BHCUs. An AI-driven tool was developed by a team of experts, comprising of healthcare design researchers and computer scientists on a university campus to automatically detect environmental safety risks in various design iterations of a BHCU during its design phase in the U.S. The unit included a milieu environment for patient observation, a nursing station, a quiet rooms, and a consultation room. To ensure the validity of the generated model and explore potential discrepancies in perception of safety between AI and users, patients and care partners’ feedback on the images was obtained through an online nationwide survey. The methodology is described in detail below:

### 2.1 Preparation of Datasets

To overcome the challenge of limited publicly available imagery of real-world BHCU environments, a large dataset of BHCU images was assembled by the team of experts using generative AI (N = 600). The assembled datasets included: 1) Images of a full-scale, low-fidelity physical mock-up of a BHCU milieu environment constructed on the university campus with cardboard, 2) Real-world images of milieu environments from various BHCU care settings across the U.S. obtained from Google search (N = 21), and 3) Synthetic images of the milieu created by the Corbu AI generative model (Schmidt, n.d.) using mock-up images and real-world references as inputs along with evidence-based, structured prompts (Figure 1). The structured prompts were designed by the team to capture critical design features (e.g., ligature-resistant features and materials) specific to mental and behavioral health environments and compatible with the existing evidence, safety codes, and regulations (Table 1).

**Figure 1.**
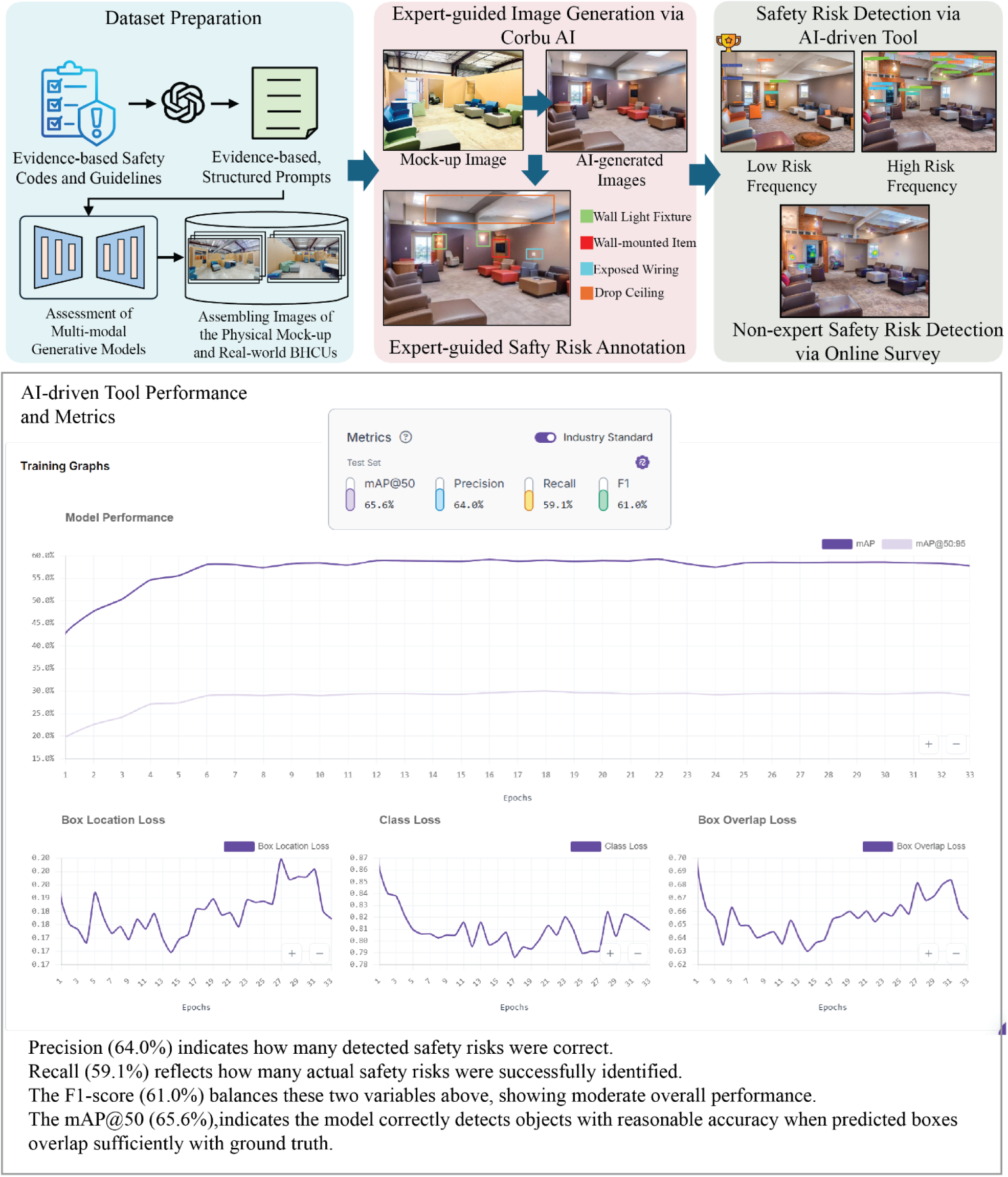
The process of dataset preparation, development and performance assessment of the AI-driven model in Roboflow

**Figure 2.**
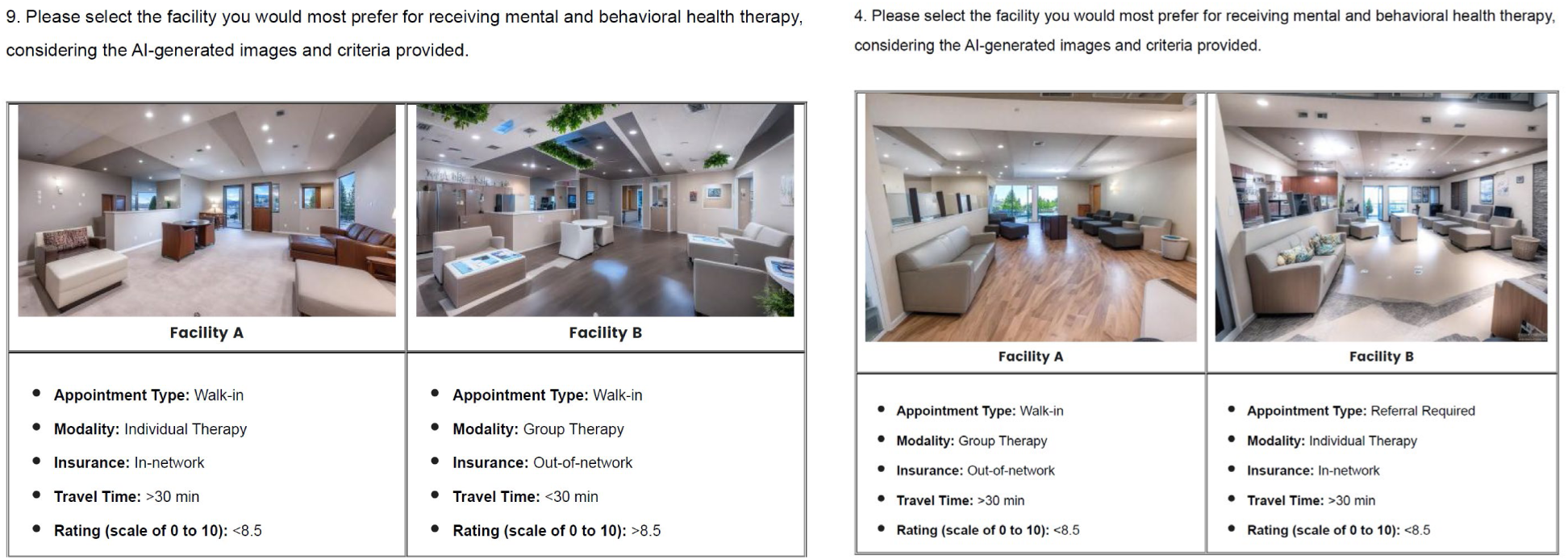
Discrete choice survey question samples

**Table 1.**
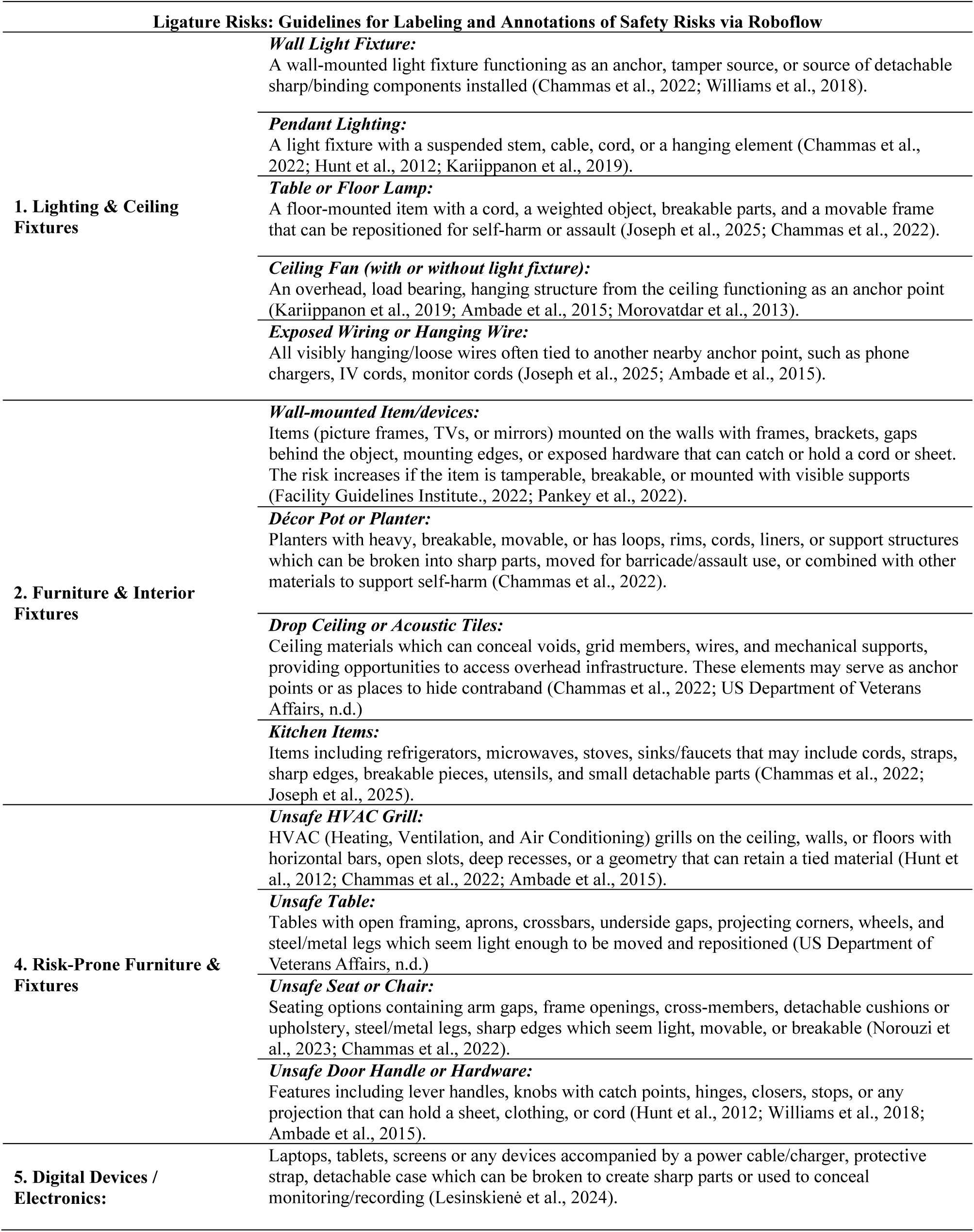
Safety guidelines considered for the AI-driven tool development.

### 2.2. Development of the AI-driven Safety Assessment Model

A selection 598 real and synthetic images (50/50 mix) were annotated using Roboflow to label target objects that were deemed unsafe with bounding boxes and class definitions. The dataset was then divided into training, validation, and testing subsets and model training was conducted using an object detection model, RF-DETR (Nano), a version of the Detection Transformer (DETR) family, adapted for efficient object detection (Robinson et al., 2025). The visual recognition model resulted in 1,424 image annotations based on the images of the BHCU environment. Metrics such as precision, recall, and Mean Average Precision (mAP) were tracked to assess how well the model performed (Figure 1).

### 2.3 Developing the Online Survey

To obtain users’ perception of safety risks based on the AI-generated images, an online survey was developed and distributed nationwide via Qualtrics platform to obtain patients’ and care partners’ feedback regarding safety risks in the BHCU images. The survey was comprised of multiple sections: 1) a discrete choice survey allowing the respondents to choose a facility to receive care based on a set of criteria, 2) heat maps for specifying environmental safety risks, 3) open-ended questions regarding the identified risks, and 4) demographics questions. The discrete choice survey was comprised of twelve question sets with each set presenting two AI-generated images of a BHCU environment with different levels of environmental safety risk frequencies along with five criteria, including care modality (individual vs. group therapy), insurance type (in-network vs. out-of-network), travel time to the facility (> 30 minutes versus < 30 minutes), and facility ratings (> 8.5 versus < 8.5 on a scale of 1 to 10) to evaluate users’ choice of care environments at the time of mental and behavioral health crisis. In section two and three of the survey, users were presented with random images from the discrete choice survey to specify environmental safety risks by clicking on images and elaborate on their perception of safety risks in images through open-ended questions. Finally, they responded to questions about themselves and their health status.

### 2.4 Sampling and Inclusion Criteria

Following a pre-test phase with 10% of the desired sample size and survey refinements, the survey was adjusted to recruit a voluntary, non-probability quota sample size of 350 participants, including patients and care partners across the U.S., to achieve 80% power, 95% confidence, and a Medium (0.3) effect size (Assele et al., 2023). Qualtrics sample services recruited participants during summer and fall 2025 based on the following inclusion criteria given: 1) Being 18 years of age or older; 2) Residing in the U.S.; 3) Having personally received care or accompanied a loved one receiving care at least once in the past 12 months for a mental or behavioral health concern in an inpatient or ambulatory or care setting, including but not limited to Behavioral Health Crisis Unit, 23-hour Crisis Observation Unit, Crisis Stabilization Unit, EmPATH Unit (Emergency Psychiatric Assessment, Treatment, and Healing), Outpatient Clinic, Emergency Department, or Urgent Care with behavioral health services. Exclusion criteria included: 1) Being under 18 years old; 2) Not residing in the U.S.; 3) Having only received care in high-security or forensic mental health units where all patients were detained under mental health or criminal legislation; and 4) Only having had brief encounters (less than ∼1 hour) such as initial triage, phone/telehealth consultations or intake screenings with the settings mentioned above.

## 3. Analytical Approach

Descriptive statistics (means, standard deviations, and percentages) and inferential statistical analysis, such as Kruskal Wallis tests were conducted to calculate and compare the frequency of environmental safety risks in real and synthetic images and pairwise comparisons between datasets. Users’ choices of facility were modeled as a function of the characteristics of the 6-choice factors using a conditional logit model. The conditional logit model is similar to logistic regression and estimates the effect of each model parameter on binary choice by seeking to maximize the likelihood function. The conditional logit model [𝜂_𝑖j_= 𝑧_j_′𝛾] indicates that utility (ηij) depends only on attributes of alternatives where 𝑧*_j_*′ represents the vector of characteristics of the j-th alternative and γ represents choice-specific parameters.

In addition to the conditional logit model, mixed multinomial conditional logit model was also utilized in this study to account for potential impacts of participant characteristics on choice behavior, including age, gender, race, education and background, annual income, overall health status, mental or emotional health status, prescribed medication intake for mental health, primary diagnosis, and encounter with mental health facilities in the past 12 months. Based on the mixed multinomial conditional logit model [𝜂_𝑖j_= 𝑥_𝑖_ ′𝛽*j* + 𝑧_𝑖j_ ′𝛾], utility (𝜂_𝑖*j*_) was modeled as a function of choice attributes and participant characteristics where 𝑥′_𝑖_ represents the characteristics that remain constant; and 𝑧′_𝑖j_ represents the participant characteristics that vary across choices.

To develop the models, forward selection method was used to identify the participant characteristics impacting choices. As the survey prompted participants to consider the hypothetical scenarios of selecting facilities where they might receive therapy during a mental or behavioral health crisis, the model was adjusted for the participant characteristics. The mixed multinomial conditional logit model fit was evaluated using likelihood-ratio (LR) tests and information criteria, including Akaike Information Criterion (AIC) and Bayesian Information Criterion (BIC). The statistical analyses and discrete choice modeling with conditional and mixed logit models were performed using the statistical package mlogit in RStudio (version 2025.09.1).

To compare the AI-driven tool versus user safety risk detection, spatial similarity analysis was conducted via Python (3.12.6) to analyze and compare heat maps generated via AI versus Qualtrics survey. The analysis included reports of the Pearson correlation coefficient (r), Structural Similarity Index Measure (SSIM) (Hayajneh et al., 2024), Jensen–Shannon value (JSD), and top-10% intersection-over-union (IoU) (Wei et al., 2024). The Pearson correlation coefficient measured the linear relationship between corresponding pixel intensities of the two sets of heat maps ranging from −1 to 1 (representing perfect agreement). SSIM assessed the perceptual similarity between two heat maps by comparing spatial patterns and structural organization of highlighted regions in images with values ranging from −1 (lower similarity) to 1 (greater similarity). The JSD value measured the shape and alignment of distributions and is used to compare probability maps in environmental and image data (Alsalama et al., 2025). The analysis also uses a top 10% IoU which assessed whether the most significant regions identified by the AI-driven tool as environmental safety risks coincided spatially with those emphasized by participants in the survey.

## 4. Results

A total of 351 participants completed the survey. The sample’s characteristics, socioeconomic, health status is demonstrated in Table 2, and are described in detail below:

**Table 2.** Participants’ demographics and clinical characteristics.

| Participant Demographics |  | Participants Clinical Characteristics |  |
| --- | --- | --- | --- |
| Age |  | Overall Health Status |  |
| 18-30 | 7.1% | Excellent | 5.7% |
| 31-45 | 30.5% | Very Good | 35.0% |
| 46-60 | 31.1% | Good | 40.2% |
| above 60 | 31.4% | Fair | 18.8% |
|  |  | Poor | 0.3% |
| Educational Background |  | Mental or Emotional Health Status |  |
| Undergraduate Degree | 35.0% | Excellent | 9.7% |
| Graduate Degree | 14.5% | Very Good | 23.4% |
| High School Degree | 18.8% | Good | 34.2% |
| Some College Education | 31.6% | Fair | 26.5% |
|  |  | Poor | 6.3% |
| Gender |  | Prescribed Medication Intake for Mental Health Conditions |  |
| Female | 62.7% | No | 33.9% |
| Male | 37.0% | Yes | 66.1% |
| Prefer not to answer | 0.3% | Primary Diagnosis for Medication Intake (If Applicable) |  |
| Race |  | Attention-deficit/hyperactivity disorder (ADHD) | 3.7% |
| Asian | 3.1% | Anxiety | 17.1% |
| Black or African American | 6.8% | Bipolar disorder | 10.0% |
| Other | 5.7% | Depression | 23.4% |
| White or Caucasian | 84.3% | None | 33.9% |
| Annual Income |  | Other | 4.3% |
| 25-99k | 55.3% | Post-traumatic stress disorder (PTSD) | 5.7% |
| 100-150k | 14.6% | Schizophrenia or other psychotic disorders | 2.0% |
| <\$25K | 17.7% | Mental Health Facility Encounter (Past 12 Months) | |
| >150k | 12.5% | Inpatient Facility | 3.42% |
| Participant Role |  | Other | 26.49% |
| Care Partner | 33.92% | Outpatient /Ambulatory Facility | 62.69% |
| Patient | 54.97% | Outpatient and Inpatient Facility | 7.41% |
| Both | 11.11% |  |  |
| Total Number of Participants: 351 |  |  |  |

**Table 3.**
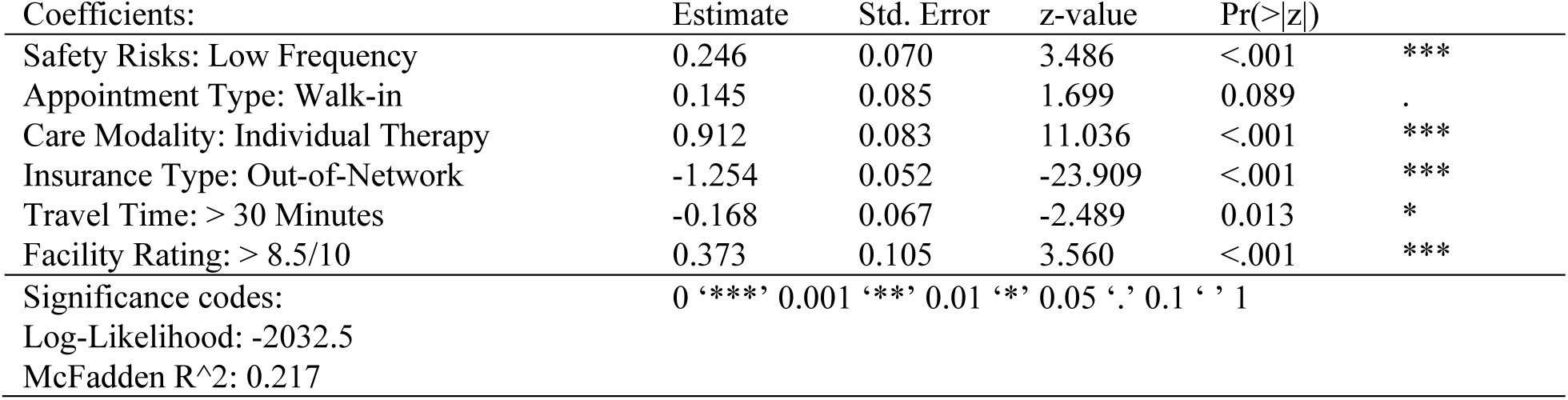
Conditional Logit Model.

***Participant Demographics:*** The majority of respondents were older adults aged above 60 years (31.4%) followed by participants aged 46–60 (31.1%) and 31–45 (30.5%). In terms of gender, the sample consisted of 220 females (62.7%), 130 males (37.0%), and 1 participant who preferred not to disclose their gender. The racial composition of the sample was predominantly White or Caucasian (84.3%) followed by Black or African American (6.8%), Asian (3.1%), and other racial groups, including American Indian or Alaska Native (5.7%). Additionally, the majority of participants in this sample held at least one college degree, including undergraduate degrees (35.0%) or graduate degrees (14.5%), while 31.6% reported some college education and 18.8% held a high school degree. The annual income levels largely fell between $25,000–$99,000 (55.3%) followed by <$25,000 (17.7%). Higher income brackets were also represented, indicating a relatively heterogeneous socioeconomic sample.

***Participants’ Clinical Characteristics:*** The majority of participants received care for a mental or behavioral health concern as patients (55.0%), accompanied a loved one who received care for a mental or behavioral health as care partners (33.9%), or both (11.1%) in the past 12 months. Most of participants rated their physical health as good (40.2%), very good (35.0%) or excellent (5.7%), while 19.08% reported fair or poor health status. Ratings of mental or emotional health status were comparatively lower among participants compared to overall health. While most participants reported good (34.2%), very good (23.4%), or excellent (9.7%), 32.8% of them rated their mental health as fair or poor, suggesting greater variability and perceived challenges in mental health relative to physical health. In addition, a substantial proportion of respondents (232 participants; 66.1%) reported that they or a loved one had taken prescription medication for a mental health condition within the past 12 months, while 119 participants (33.9%) who reported no such use. Among reported conditions, depression (23.4%), anxiety (17.1%), and bipolar disorder (10.0%) accounted for main primary conditions associated with medication use. A relatively large proportion of respondents (33.9%) did not specify a condition.

### 4.1 Conditional Logit Model

Results presented in Table 3 demonstrate the conditional logit (alternative-specific) model where no individual-specific variables were considered. The coefficients in Table 3 describe how the utility of each alternative, including Facility A (selection proportion = 69.3%) and Facility B (selection proportion = 30.7%), changed as its attributes changed. Based on the model, frequency of environmental safety risks in AI-generated facility images, care modality (individual vs. group therapy), insurance type (in-network vs. out-of-network), travel time to the facility (> 30 minutes versus < 30 minutes), and facility ratings (> 8.5 versus < 8.5 on a scale of 1 to 10) were attributes that significantly affected the selection of alternatives, Facility A versus Facility B.

**Table 3.**
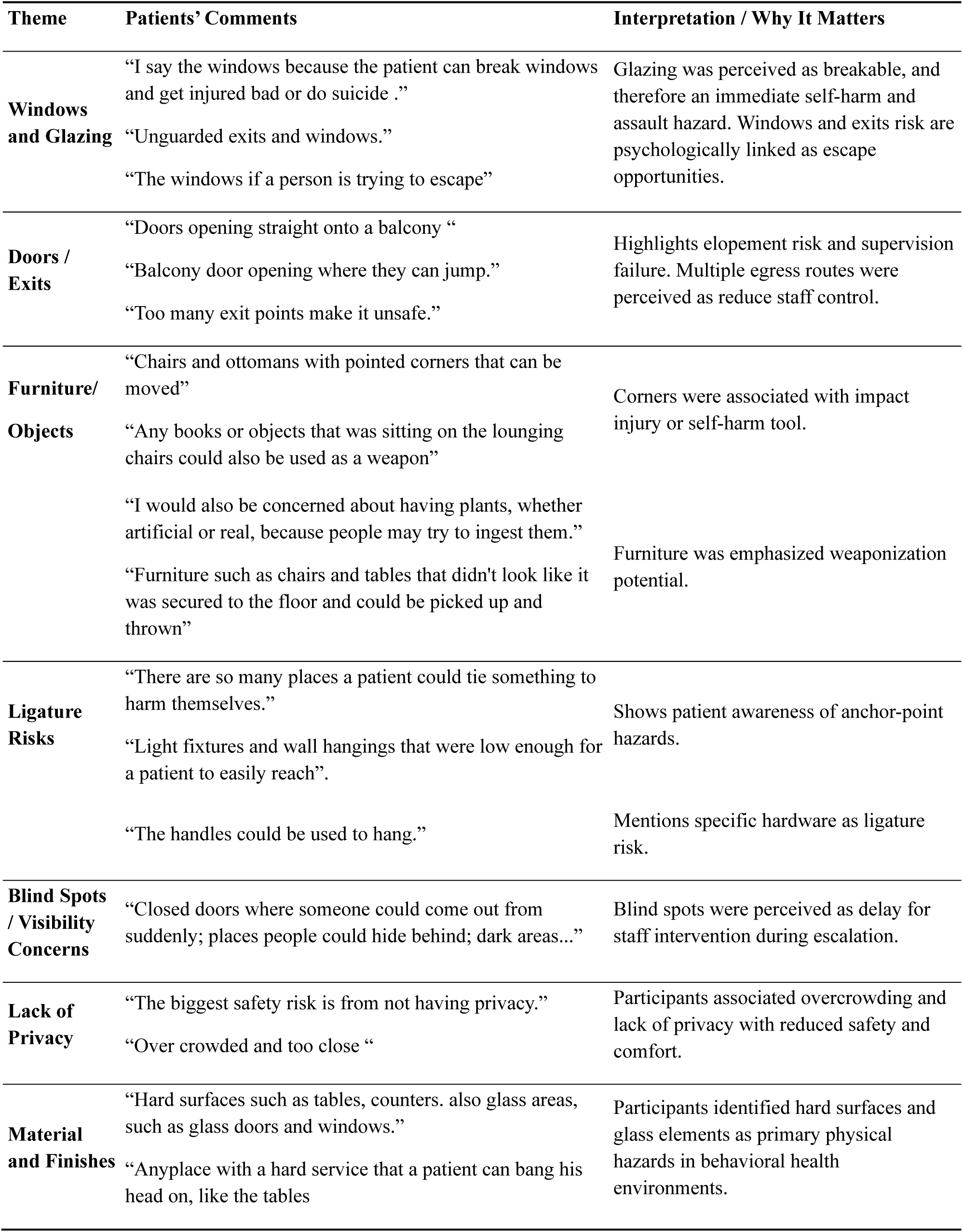
Participants’ feedback regarding safety risks identified in AI-generated unit images.

Results indicated that when alternatives had images of facilities with lower frequency of environmental safety risks, the odds of choosing that alternative increased by 28%, holding all other attributes constant. Given (p < .001, OR ≈ 1.28), participants significantly preferred the facility options showing lower safety risks. Following safety risks, an alternative with individual care modality option had about 2.5 times the odds of being chosen compared to the group therapy modality option, holding everything else constant, showing that participants strongly preferred individual therapy options at the time of crisis (p < .001, OR ≈ 2.49). Similarly, the odds of choosing an alternative with out-of-network insurance option was about 71% lower than choosing an alternative with an in-network option. This large negative effect showed that participants strongly avoided out-of-network options, even when other attributes (risks, modality, rating, etc.) were favorable (p < .001, OR ≈ 0.29). In terms of travel time to facilities, a travel times longer than 30 minutes to a facility at the time of crisis reduced the odds of choosing that alternative by 15% among participants. While this reveals that participants disliked extra travel time, the effect size was moderately small compared to other attributes, including insurance type and care modality (p = 0.013, OR ≈ 0.85). Moreover, results showed that alternatives offering walk-in appointments had about 16% higher odds of being chosen by participants than those accepting referrals only, all else equal. While the impact of this attribute was marginal (p = 0.089, OR ≈ 1.16), it still suggests that participants preferred walk-in appointments for mental and behavioral facilities, such as BHCUs, at the time of crisis. Finally, results indicated that alternatives with facility ratings higher than 8.5 on a scale of 1 to 10 increased odds of being chosen by 45% among participants. In other words, higher rating for a facility was a strong positive driver of participants’ choice (p < .001, OR ≈ 1.45), stronger than travel time and appointment type, but smaller than care modality and insurance type in absolute impact.

### 4.2 Mixed Multinomial Conditional Logit Model

Based on Table 4, results indicated no inherent preference for either Facility A or B once participant characteristics were included in the model. While there were instances of AI-generated images with environmental safety risks in both alternatives, the median environmental safety risk counts detected by the AI-driven tool for images presented in alternative “Facility B” (Median = 5, MAD = 2) was slightly higher than “Facility A” (Median = 3.5, MAD = 1.5) in this survey across the choice sets. As illustrated in Table 4, attributes, including appointment type, care modality, insurance type, travel time to facility, and facility rating were significantly associated with participant choices. In addition, participant characteristics, such as mental and overall health status, race, age, prior interaction type with mental and behavioral health facilities, and type of mental and behavioral health diagnosis seemed to have impacted choices as well.

**Table 4.** Mixed Multinomial Conditional Logit Model accounting for participants’ characteristics.

| Coefficients: | Estimate | Std. Error | z-value | Pr(>\|z\|) |  |
| --- | --- | --- | --- | --- | --- |
| (Intercept): Facility B | 0.300 | 0.317 | 0.948 | 0.343 |  |
| Safety Risks: Low Frequency | 0.119 | 0.077 | 1.542 | 0.123 |  |
| Appointment: Walk-in | -0.379 | 0.110 | -3.449 | 0.001 | *** |
| Care Modality: Individual Therapy | 0.801 | 0.087 | 9.264 | <.001 | *** |
| Insurance: Out-of-network | -1.087 | 0.059 | -18.305 | <.001 | *** |
| Travel Time: > 30 minutes | -0.247 | 0.069 | -3.587 | <.001 | *** |
| Facility Rating: > 8.5/10 | 0.401 | 0.107 | 3.744 | <.001 | *** |
| Mental Health Status: Fair | -0.126 | 0.171 | -0.734 | 0.463 |  |
| Mental Health Status: Good | 0.112 | 0.157 | 0.708 | 0.479 |  |
| Mental Health Status: Poor | -0.134 | 0.228 | -0.587 | 0.557 |  |
| Mental Health Status: Very Good | -0.432 | 0.161 | -2.687 | 0.007 | ** |
| Facility Encounter Type: Other | -0.429 | 0.224 | -1.920 | 0.055 | . |
| Facility Encounter Type: Outpatient | -0.501 | 0.216 | -2.316 | 0.021 | * |
| Facility Encounter Type: Outpatient + Inpatient | -0.438 | 0.253 | -1.729 | 0.084 | . |
| Overall Health Status: Fair or Poor | -0.799 | 0.215 | -3.717 | <.001 | *** |
| Overall Health Status: Good | -0.587 | 0.195 | -3.005 | 0.003 | ** |
| Overall Health Status: Very Good | -0.408 | 0.185 | -2.202 | 0.028 | * |
| Race: Asian | -0.584 | 0.255 | -2.291 | 0.022 | * |
| Race: Black or African American | 0.146 | 0.163 | 0.900 | 0.368 |  |
| Race: Other | 0.448 | 0.168 | 2.659 | 0.008 | ** |
| Age: 31-45 | 0.501 | 0.171 | 2.932 | 0.003 | ** |
| Age: 46-60 | 0.055 | 0.173 | 0.317 | 0.751 |  |
| Age: above 60 | 0.223 | 0.176 | 1.267 | 0.205 |  |

Significance codes: 0 '\*\*\*' 0.001 '\*\*' 0.01 '\*' 0.05 '.' 0.1 ' ' 1 Log-Likelihood: -1958.3 McFadden R<sup>2</sup>: 0.246 Likelihood ratio test: chisq = 1280.5 (p value = <.001) AIC: 3962.645 and BIC: 23

Results revealed that for participants indicating a very good level of mental or emotional health in general in this survey, there was about 35% lower odds of choosing Facility B options (p = 0.007, OR ≈ 0.65) with slightly higher counts of safety risks than Facility A. Also, those respondents who evaluated their overall health status “Fair or Poor” (p < .001, OR ≈ 0.45) were significantly less likely to choose Facility B. Additionally, results showed that respondents who only interacted with outpatient facilities, including, emergency departments, urgent care, or behavioral health crisis units were also significantly less likely to choose B (about 39% lower odds) than those with an inpatient hospitalization experience in the past 12 months (p = 0.021, OR ≈ 0.61). Among participants with different races, Asian respondents had about 44% lower odds of choosing Facility B (p = 0.022, OR ≈ 0.56) while others, including American Indian, Alaska Native, or Native Hawaiian had about 57% higher odds of choosing Facility B (p = 0.008, OR ≈ 1.57) compared to White or Caucasian respondents. Moreover, respondents aged 31–45 had about 65% higher odds of choosing Facility B than the youngest age group (18 to 30); however, no significant differences for respondents aged 46–60 or >60 were observed.

### 4.3 Detection of Environmental Safety Risks Based on Images: Patients and Care Partners versus the AI-driven Tool

Following the discrete choice survey, participants were asked to identify and mark areas or features that could present risks to patient safety by clicking on each AI-generated image presented to them in the discrete choice survey. The same images were also analyzed by the AI-driven tool developed by the team of experts to automatically detect and mark environmental safety risks. The survey results versus AI-driven tool’s output are illustrated in Figure 3 and 4.

**Figure 3.**
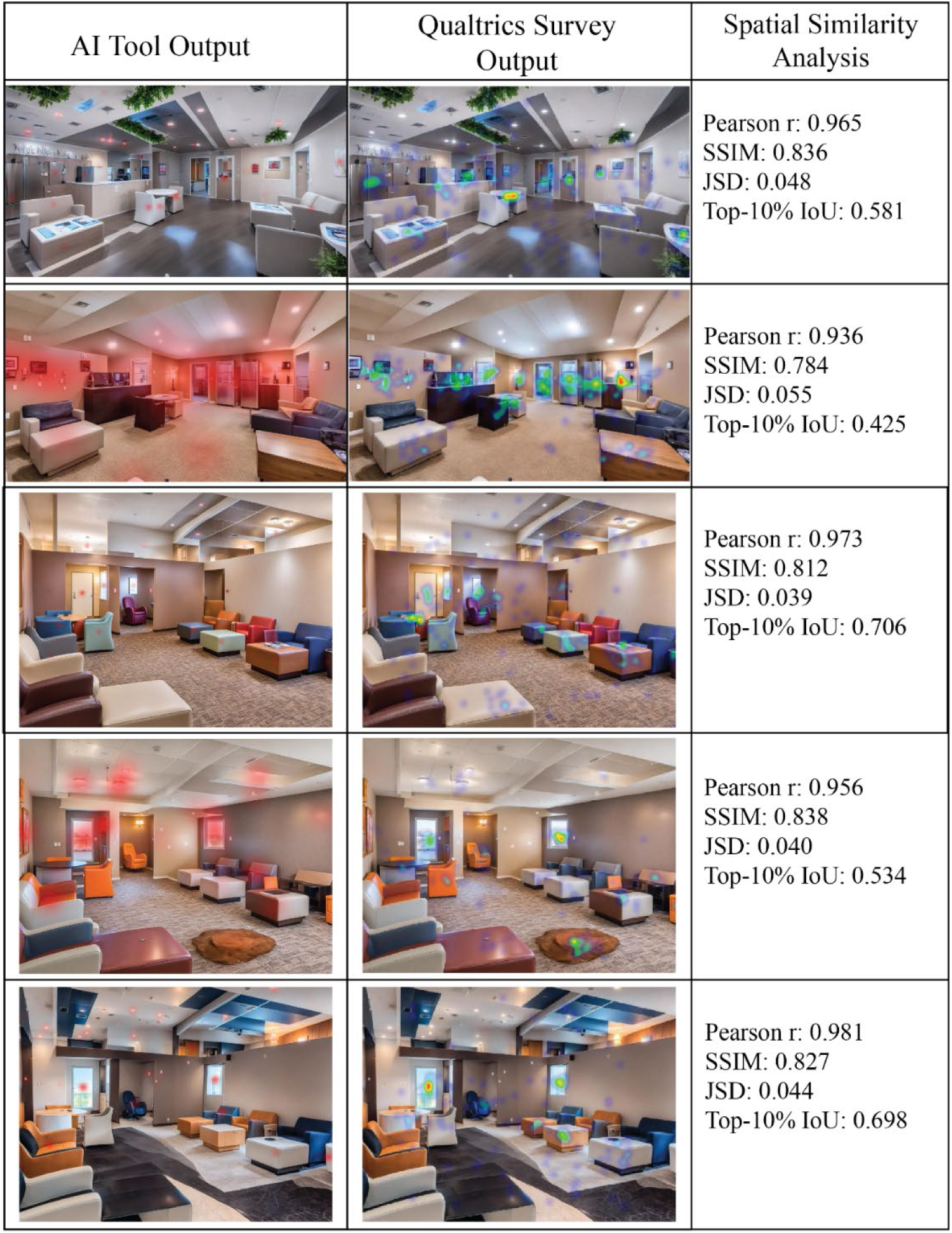
Comparisons of the heat maps obtained from safety risk analysis of the images by the AI-driven tool versus survey participants

**Figure 4.**
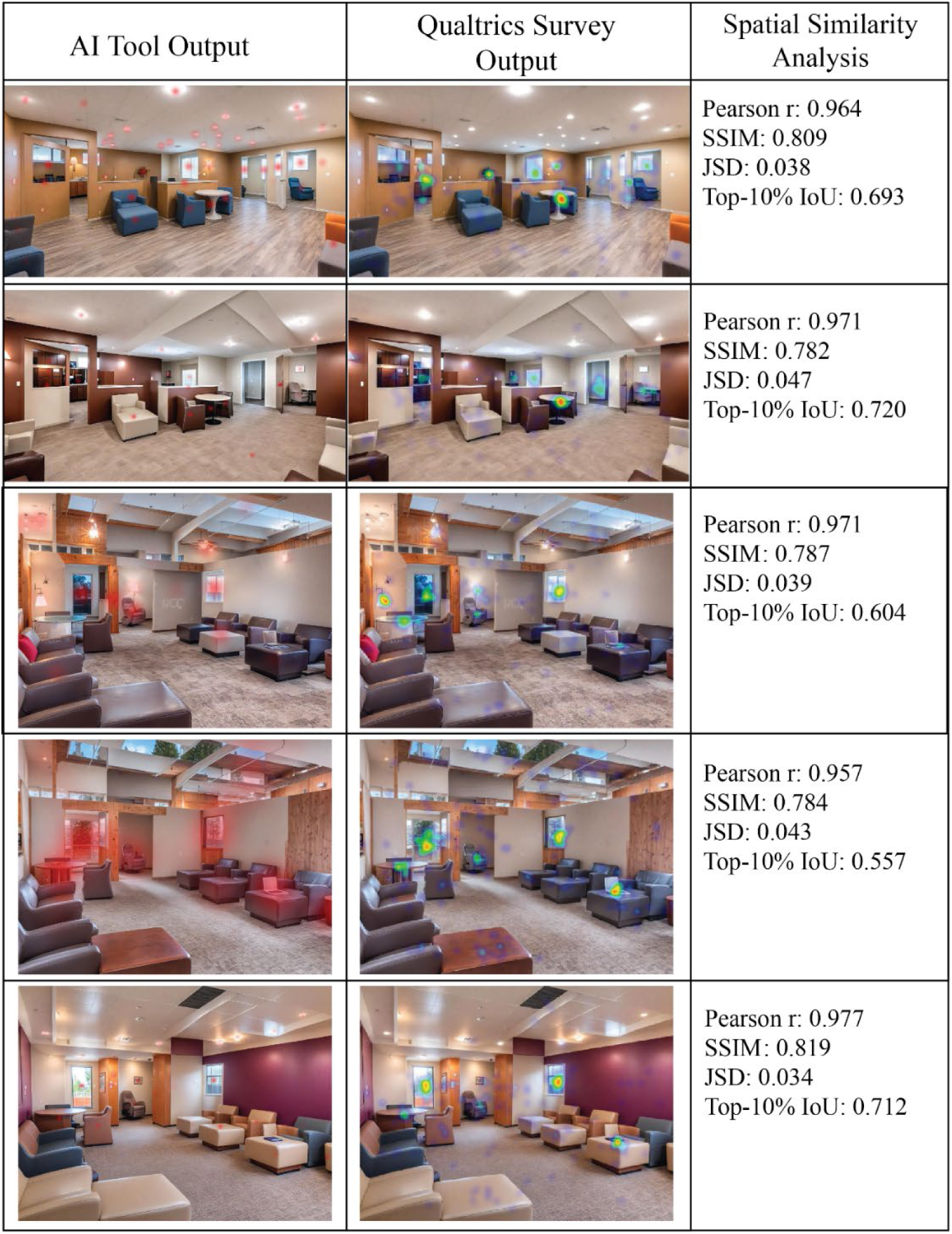
Comparisons of the heat maps obtained from safety risk analysis of the images by the AI-driven tool versus survey participants

The analysis revealed a high degree of agreement between AI-generated heat maps and human-reported heat maps derived from the Qualtrics survey. A strong positive Pearson correlation ranging from (r = 0.94) to (r = 0.98) indicates that regions with higher intensity in the AI heat map significantly corresponded to regions marked by participants in the survey heat maps. Structural similarity analysis further supported this finding, with a relatively high SSIM values ranging from 0.78 to 0.84, suggesting substantial alignment in the overall spatial structure and distribution of highlighted areas of the heat maps. Distributional comparison using JSD also revealed low values ranging from 0.03 to 0.05, indicating that the two sets of heatmaps shared very similar spatial probability distributions. In addition, analysis of hotspot overlap demonstrated meaningful spatial convergence, with a top-10% IoU of 0.43 to 0.72, showing that the most salient regions identified by the AI-driven tool as environmental safety risks substantially overlapped with those identified by participants in the survey heatmaps.

Following identifying safety risks, the survey asked participants to elaborate on specific areas or features that seemed to pose the highest environmental safety risks to patients in the images. Several themes were extracted based on the participants responses to the open-ended questions in this part of the survey, including ligature risks, windows and glazing, doors and exit points, furniture pieces, blind spots and visibility concerns, privacy concerns, and materials or finishes. Comparing heatmaps created via AI versus participants’ input revealed that participants seemed to consider glazing a safety risk in doors, windows, or even nursing station areas more frequently than the AI tool stating, “*the patient could break windows and get injured bad or do suicide*”. Other comments included “*Light fixtures and wall hangings that were low enough for a patient to easily reach*” or “*I would also be concerned about having plants, whether artificial or real, because people may try to ingest them*”. Participants also marked unidentified objects or furniture features explaining “*anywhere the furniture sticks out and also the rug on the floor is a risk!”.* A summary of the extracted theme and participants’ comments are presented in Table 3.

### 4.4 Discussion

This study investigated differences in the detection and perception of environmental safety risks between AI and end-users in BHCU environments through a nationwide survey in the U.S. Findings based on the discrete choice survey revealed that between alternatives “Facility A” and “Facility B” with images presenting varying levels of environmental safety risks (low versus high), participants significantly preferred the options with a facility image demonstrating lower frequency of environmental safety risks (e.g., ligature points). The model also revealed that attributes, including care modality, insurance type, travel time to facility and facility ratings were significantly associated with participants’ choices of alternatives. However, comparing Odds Ratios associated with each attribute showed that users’ choices of facility were strongly shaped by in-network insurance coverage and care modality (individual therapy options) rather than environmental safety risks. The significant impacts of in-network insurance coverage, closer travel distance from home and facility ratings on patients’ choices of healthcare facilities have been also shown in previous studies. A study of 200 patients through a discrete choice experiment for outpatient facilities also indicated that hospital rating and travel time significantly affected patients’ choices of a facility for joint arthroplasty procedures (Schwartz et al., 2021). Only one study investigating patients’ hospital choice through a nationwide discrete choice survey of 652 patients revealed that design features in healthcare environments, such as window design, also significantly impacted patients’ choice of the facility (Woo et al., 2022). Nevertheless, none of these studies explored patients’ preferences for mental and behavioral health environments or their perception of safety risks as a driver of facility choice. While the present study strived to address this gap, findings showed that the effect of image-based environmental safety risks on facility selections was only statistically significant when no individual-specific characteristics related to participants were considered. When specific characteristics, such as health status, type of encounter with mental health facilities, race, and age, were included in the model, the effect of this attribute decreased while other attributes, including insurance, travel distance, care modality, facility rating, and appointment type remained significantly associated with participants’ choices of alternatives.

When comparing detection of safety risks between AI and participants (patients and care partners) based on synthetic BHCU images, findings indicated a strong spatial alignment between AI-driven and human-reported heatmaps for environmental safety risk detection. This finding aligns with prior studies in healthcare settings demonstrating that AI-driven assistive tools could achieve performance comparable to—or exceeding—that of human observers in detecting safety risks associated with adherence to hospital protocols (Kim et al., 2025; Singh et al., 2020). Despite a high degree of alignment between safety risk detection heatmaps, the perception of safety risks—defined as the rationale guiding the selection of specific risk features in images—was not fully aligned between the AI-driven tool and non-expert respondents, highlighting limitations of assistive AI-driven tools in environmental safety risk detection and assessment. For instance, windows and glazing were perceived as safety risks by participants more frequently as they were psychologically linked to elopement risks or opportunities for injury to self or others through broken glass. Nevertheless, the AI-driven tool was mainly programmed by experts to detect ligature risks related to certain window features and hardware rather than the glazing as the tool could not determine the type of glazing (e.g., tamper-resistant versus regular) based on the image-based input data. As safety risks related to materials and finishes, such as glazing, constitute a large category of risks in mental and behavioral health environments (Facility Guidelines Institute, 2022), such limitation might prevent AI-driven assistive tools to detect the full range of safety risks in such environments. Additionally, survey participants marked random or unfamiliar objects in synthetic images as potential safety risks while the AI-driven tool was unable to detect or flag items that were unknown or not previously labeled by experts. The existing literature also emphasizes that detection of out-of-label safety risks is a persisting challenge for many AI-driven, computer vision systems as they fall outside the training distribution (Zeng, 2025). Considering the various types of behavioral crisis care environments across the country providing care for patients, larger datasets would be required to properly train the assistive AI-driven tools in identifying safety risks for non-experts.

Other limitations of this study relate to the fact that the majority of participants in this nationwide survey were older adults. Additional studies are required to investigate perception of safety among young adults or adolescents and explore similarities and differences in their perception versus AI-driven tools. Additionally, the survey incorporated multiple AI-generated images of a single BHCU, derived from a full-scale physical mock-up, along with a limited number of real-world BHCU images to support training of the AI-driven model, given the limited publicly available imagery of such environments. Future research can build on the existing research by using larger datasets with images of real-world mental and behavioral health units to increase the validity and generalizability of the study. Moreover, additional research is required to elucidate how health status can impact perception of safety and facility choice among mental and behavioral health patients and their care partners in discrete choice surveys.

### 4.5 Conclusion

This study incorporated a nationwide discrete choice survey and revealed that presence of environmental safety risks—particularly ligature points— in images of mental and behavioral health facilities, such as BHCUs, significantly influenced patients’ and care partners’ selection of care environments. While other factors, including insurance type, care modality (individual versus group therapy), and travel time remained significant drivers of users’ choice of facility, findings showed that decisions were also shaped by users’ safety perception of the care environment. In addition, findings indicated strong alignments between the frequency of user-identified environmental safety risks and AI-driven detection of risks through heatmaps. However, discrepancies existed between the two groups in how risks were perceived and identified. While the overlap suggests that pre-trained AI-driven tools can effectively capture a range of visible environmental safety risks, limitations still exist in understanding certain risks related to the type of materials (e.g., type of grazing) or detecting unknown, out-of-label safety risks in facility images. Future research can address these limitations using larger datasets and broader range of data from both non-expert users and AI-driven, expert-informed systems to improve the performance of such assistive tools and support safer and more responsive mental and behavioral health environments.

## Declaration of Conflicting Interests

The Authors declare that there is no conflict of interest.

## Data Availability

All data produced in the present study are available upon reasonable request to the authors.

## Acknowledgement

The author would like to thank the Texas A&M College of Architecture and Qualtrics for their contributions to this study. Also, the author would like to thank PhD students, Hani Patel and Elnaz Shadkami from Texas A&M University, Department of Architecture, for their contributions to data sorting and labeling in this study.

## Funding

This work was supported by The College of Architecture at Texas A&M University.

## Ethical Considerations

Ethical approval for this study was obtained from The Texas A&M University Institutional Review Board, STUDY2024-0920.

“Not Peer-reviewed” disclaimer

This manuscript is a preprint and has not been peer-reviewed. It should not be used to guide clinical practice.

## Notes

### Competing Interest Statement

The authors have declared no competing interest.

